# Qualitative forecast and temporal evolution of the disease spreading using a simplified model and COVID-19 data for Italy

**DOI:** 10.1101/2020.06.22.20137133

**Authors:** Roberto Simeone

**Affiliations:** None

**Keywords:** COVID-19, Italy, SEIR, compartmental model, reproduction number

## Abstract

In a previous paper [1] a simplified SEIR model applied to COVID-19 cases detected in Italy, including the lockdown period, has shown a good fitting to the time evolution of the disease during the observed period.

In this paper that model is applied to the initial data available for Italy in order to forecast, in a qualitative way, the time evolution of the disease spreading. The values obtained are to be considered indicative.

The same model has been applied both to the data relating to Italy and to some italian regions generally finding good qualitative results.

The only tuning parameter in the model is the ‘incubation period’ *τ*.

In this modelization the tuning parameter, together with the calculated growth rate of the exponential curve used to approximate the early stage data, are in strong relationship with the compartments’ transfer rates.

The relationships between the parameters simplify modeling by allowing a rough (not supported by statistical considerations) forecast of the time evolution, starting from the first period of growth of the diffusion.

**Conclusions:** A simplified compartmental model that uses only the incubation period and the exponential growth rate as parameters is applied to the COVID-19 data for Italy in several periods of the initial growth of the diffusion showing the different stages of the spread evolution. The simplification is based on the strong protection measures that were in place in Italy during the lockdown period after the initial free diffusion.

## Revision history

**Revision # 1** Errata corrige in the system differential equation 1: in the the derivative of S were reported a wrong additional term N. Now the equation 1 is correct.

**Revision #2** A new approach is used to detect the exponential rate and new concept for the transfer coefficients. The nrw approach is described in [1] and is summarized below.

- Exponential rate: The old criteria was oriented to the growth of the cases: *y_Δt_* = *y_0_* * *e^kΔt^* thus: *y_0_* + Δ*y* = *y_0_* * *e^kΔt^*. The exponential growth rate was then: *k* = *log(*1 + Δ*y/y_0_)/*Δ*t*. The new criteria is oriented to the growth of the differences Ay = *e^kΔt^ −* 1 obtaining: *k* = log(1 + Δ*y*)/Δ*t*.
- **Transfer coefficients:** The new approach is based on the following assumptions: *α_SE_* = *ke^kδ^* [day^−1^]: this coefficent is supposed to be the variation of the exponential growth per unit of time (*δ* = 1 *day)*. *α_EI_* = 1/*τ* [day^-1^] where *τ* is the incubation period (this assumption is not changed). *α_IT_* = *kδ/τ* [day^−1^] this coefficent is supposed to be proportional to the ratio *kδ/τ*. The constant *δ* =1 [day] represent the unit variation in time.
- **Basic reproduction number:** With the above assumptions, the basic reproduction number become: 𝓡_0_ = *α_SE_*/*α_IT_* = *e^kδ^τ/δ* [adimensional] (δ = 1 day)
- **Revision # 2 summary:** The old approach, although adapting well to the data, presented several inconsistencies in the parameters, and in particular on the relationship between 𝓡_0_ and *k*. In this revision the new approach still shows a good fit to the data and shows congruent relationships between the parameters.

## 1 Introduction

A simplified SEIR compartmental model seems to fit quite well the evolution of COVID-19 during the period of social isolation in Italy [1]. In this paper the model is used to forecast the diffusion behavior starting from the initial stage of infection growth.

The dataset is the same of the previous paper [1] and is available on the website [2] managed by italian government. The model used is a simplification of the Susceptible-Exposed-Infected-Recovered type (see [3], [5], [4]) and the simplification is based on the relationships between the model parameters and the growth rate of the exponential curve in the early stage of diffusion (see [1]). The dataset cover a period where after a initial growth of the COVID-19 diffusion a ‘lockdown’ period, characterized by strong social isolation rules, were imposed to act effectively against the rapid spread of the disease.

The lockdown period, simplifying, can also be thought of as a type of treatment for infective individuals. We can therefore imagine that infective individuals, detected daily, in general are managed (’treated’) in such a way that they cannot infect other susceptible individuals. This ‘treatment’ includes both hospitalization and isolation. To avoid confusing the SEIR model with this simplification, in this context, we will call T (Treated) the compartment of infected people who are no longer able to transmit the pathogen.

The social isolation, also allows us to neglect any potential transfers between the various departments beyond the required path *S → E → I → T*. The ‘treated’ compartment in this case can be seen as a sample of individuals in whom the disease has been widespread and the spread is characterized by the model coefficients. The significance of the sample is related to the number of detections of the disease, and their distribution, carried out in the population.

The model can be set up easily once initial the growth rate is calculated and a suitable value for the incubation period is defined. In this work the data used are normalized to 1000 individuals in order to generalize the results. The first part of data, in the growing period, are used to calculate the growth rate k, the remaining data are used for comparison with the model’s forecasting.

## 2 Conclusions

A simplified compartmental model [1], when strong protection measures are put in place after the initial free diffusion, can be used to easily predict a qualitative behavior of a disease diffusion starting from the early stage. It is obviously necessary that the number of detections of the disease and their distribution, carried out in the population, are statistically significant, in this case the simplified model can be used to roughly estimate the qualitative behavior of the spread.

The only tuning parameter in the simplified model is the ‘incubation period’ *τ*. In this modelization the tuning parameter, together with the calculated growth rate *κ* of the exponential curve used to approximate the early stage data, are in strong relationship with the compartments’ transfer rates. The relationships between the parameters simplify modeling by allowing a rough (not supported by statistical considerations) forecast of the time evolution, starting from the first period of growth of the diffusion.

## 3 Model parameters and detection criteria

The key parameters needed to set up a simplified SEIT model are (see [1]):

- the incubation period *τ*;
- the exponential growth rate *k* calculated in the initial diffusion. In this case we are oriented to the growth of the differences Δ*y* = *e^kΔt^ −* 1 obtaining: *k* = *log*(1 + Δ*y)/*Δ*t*.

It is assumed that the number, the quality and the distribution of tests to detect the disease in the population are statistically significant; in this case, using the simplified SEIT model it is possible to easily characterize, in a qualitative way, the disease diffusion in the compartments S,E,I,T over time.

The system of differential equations for the simplified model *S → E → I → T* is:

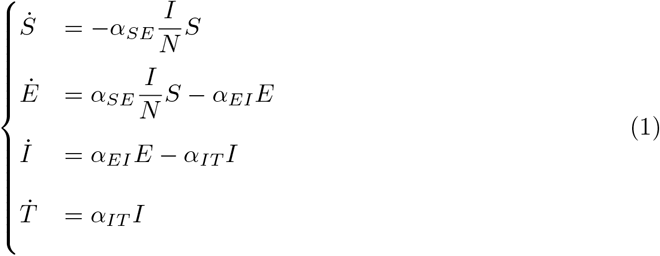

where N is the total number of individuals: *S* + *E* + *I* + *T* The temporal evolution follows a disruption of the equilibrium at *t* = 0:

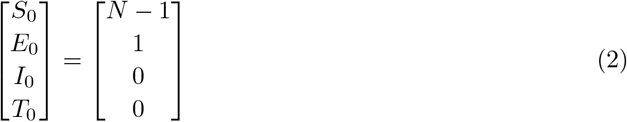

At *t* = 0 the number of individuals susceptible to the disease is *S*_0_ = *N −* 1 and there is one individual exposed. In this work the data are normalized to 1000 individuals.

In this simplification it is assumed that there is a unique available path *S* → *E* → *I* → *T*. In this case, following the assumptions described in (see [1]) the transfer coefficients between compartments are:

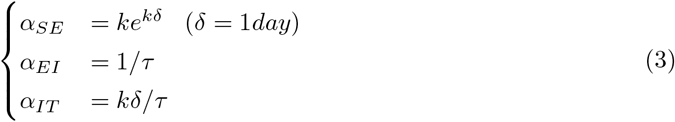

The basic reproduction number calculated as spectral radius of the next generation matrix (see [3], [4], [5]) in the simplified model (see [1]) is:

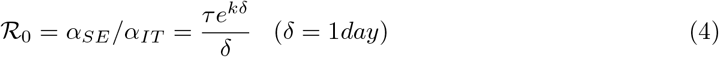

The criteria used in this context to find *k* is that a new *k* is calculated when the daily variation 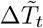 of the detected cases exceed the last maximum of daily variation.

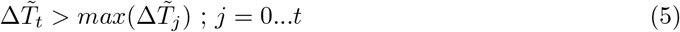

In this case a new *k* is calculated as:

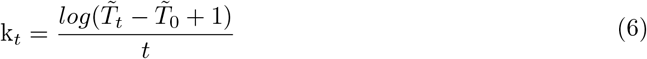

where 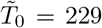 individuals at *t* = 0 corresponding to the first day of detection 2020-02-24 (see table 2).

**Table 1:** Records corresponding to the days when exceeding the last maximum daily variation value (the first 5 days are not used to detect k). The growth rate k is calculated as per (6). The full dataset is in tab. 2

| t | Day | $\tilde{T}_t$ | $\Delta\tilde{T}_t$ | $k$ |
| --- | --- | --- | --- | --- |
| 6 | 2020-03-01 | 1694 | 566 | 1.215 |
| 9 | 2020-03-04 | 3089 | 587 | 0.884 |
| 10 | 2020-03-05 | 3858 | 769 | 0.820 |
| 11 | 2020-03-06 | 4636 | 778 | 0.763 |
| 12 | 2020-03-07 | 5883 | 1247 | 0.720 |
| 13 | 2020-03-08 | 7375 | 1492 | 0.683 |
| 14 | 2020-03-09 | 9172 | 1797 | 0.650 |
| 16 | 2020-03-11 | 12462 | 2313 | 0.588 |
| 17 | 2020-03-12 | 15113 | 2651 | 0.565 |
| 19 | 2020-03-14 | 21157 | 3497 | 0.524 |
| 20 | 2020-03-15 | 24747 | 3590 | 0.505 |
| 23 | 2020-03-18 | 35713 | 4207 | 0.456 |
| 24 | 2020-03-19 | 41035 | 5322 | 0.442 |
| 25 | 2020-03-20 | 47021 | 5986 | 0.430 |
| 26 | 2020-03-21 | 53578 | 6557 | 0.419 |

**Table 2:** Dataset of detected cases 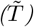 and daily variations 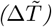 for Italy extracted from [2]

| t | Day | $\tilde{T}_t$ | $\Delta\tilde{T}_t$ | t | Day | $\tilde{T}_t$ | $\Delta\tilde{T}_t$ |
| --- | --- | --- | --- | --- | --- | --- | --- |
| 0 | 2020-02-24 | 229 | 0 | 15 | 2020-03-10 | 10149 | 977 |
| 1 | 2020-02-25 | 322 | 93 | 16 | 2020-03-11 | 12462 | 2313 |
| 2 | 2020-02-26 | 400 | 78 | 17 | 2020-03-12 | 15113 | 2651 |
| 3 | 2020-02-27 | 650 | 250 | 18 | 2020-03-13 | 17660 | 2547 |
| 4 | 2020-02-28 | 888 | 238 | 19 | 2020-03-14 | 21157 | 3497 |
| 5 | 2020-02-29 | 1128 | 240 | 20 | 2020-03-15 | 24747 | 3590 |
| 6 | 2020-03-01 | 1694 | 566 | 21 | 2020-03-16 | 27980 | 3233 |
| 7 | 2020-03-02 | 2036 | 342 | 22 | 2020-03-17 | 31506 | 3526 |
| 8 | 2020-03-03 | 2502 | 466 | 23 | 2020-03-18 | 35713 | 4207 |
| 9 | 2020-03-04 | 3089 | 587 | 24 | 2020-03-19 | 41035 | 5322 |
| 10 | 2020-03-05 | 3858 | 769 | 25 | 2020-03-20 | 47021 | 5986 |
| 11 | 2020-03-06 | 4636 | 778 | 26 | 2020-03-21 | 53578 | 6557 |
| 12 | 2020-03-07 | 5883 | 1247 | 27 | 2020-03-22 | 59138 | 5560 |
| 13 | 2020-03-08 | 7375 | 1492 | 28 | 2020-03-23 | 63927 | 4789 |
| 14 | 2020-03-09 | 9172 | 1797 | 29 | 2020-03-24 | 69176 | 5249 |

| t | Day | $\tilde{T}_t$ | $\Delta\tilde{T}_t$ |
| --- | --- | --- | --- |
| 30 | 2020-03-25 | 74386 | 5210 |
| 31 | 2020-03-26 | 80539 | 6153 |
| 32 | 2020-03-27 | 86498 | 5959 |
| 33 | 2020-03-28 | 92472 | 5974 |
| 34 | 2020-03-29 | 97689 | 5217 |
| 35 | 2020-03-30 | 101739 | 4050 |
| 36 | 2020-03-31 | 105792 | 4053 |
| 37 | 2020-04-01 | 110574 | 4782 |
| 38 | 2020-04-02 | 115242 | 4668 |
| 39 | 2020-04-03 | 119827 | 4585 |
| 40 | 2020-04-04 | 124632 | 4805 |
| 41 | 2020-04-05 | 128948 | 4316 |
| 42 | 2020-04-06 | 132547 | 3599 |
| 43 | 2020-04-07 | 135586 | 3039 |
| 44 | 2020-04-08 | 139422 | 3836 |
| 45 | 2020-04-09 | 143626 | 4204 |
| 46 | 2020-04-10 | 147577 | 3951 |
| 47 | 2020-04-11 | 152271 | 4694 |
| 48 | 2020-04-12 | 156363 | 4092 |
| 49 | 2020-04-13 | 159516 | 3153 |
| 50 | 2020-04-14 | 162488 | 2972 |
| 51 | 2020-04-15 | 165155 | 2667 |
| 52 | 2020-04-16 | 168941 | 3786 |
| 53 | 2020-04-17 | 172434 | 3493 |
| 54 | 2020-04-18 | 175925 | 3491 |
| 55 | 2020-04-19 | 178972 | 3047 |
| 56 | 2020-04-20 | 181228 | 2256 |
| 57 | 2020-04-21 | 183957 | 2729 |
| 58 | 2020-04-22 | 187327 | 3370 |
| 59 | 2020-04-23 | 189973 | 2646 |
| 60 | 2020-04-24 | 192994 | 3021 |
| 61 | 2020-04-25 | 195351 | 2357 |
| 62 | 2020-04-26 | 197675 | 2324 |
| 63 | 2020-04-27 | 199414 | 1739 |
| 64 | 2020-04-28 | 201505 | 2091 |
| 65 | 2020-04-29 | 203591 | 2086 |
| 66 | 2020-04-30 | 205463 | 1872 |
| 67 | 2020-05-01 | 207428 | 1965 |
| 68 | 2020-05-02 | 209328 | 1900 |
| 69 | 2020-05-03 | 210717 | 1389 |
| 70 | 2020-05-04 | 211938 | 1221 |
| 71 | 2020-05-05 | 213013 | 1075 |
| 72 | 2020-05-06 | 214457 | 1444 |
| 73 | 2020-05-07 | 215858 | 1401 |
| 74 | 2020-05-08 | 217185 | 1327 |
| 75 | 2020-05-09 | 218268 | 1083 |
| 76 | 2020-05-10 | 219070 | 802 |
| 77 | 2020-05-11 | 219814 | 744 |
| 78 | 2020-05-12 | 221216 | 1402 |
| 79 | 2020-05-13 | 222104 | 888 |
| 80 | 2020-05-14 | 223096 | 992 |
| 81 | 2020-05-15 | 223885 | 789 |
| 82 | 2020-05-16 | 224760 | 875 |
| 83 | 2020-05-17 | 225435 | 675 |
| 84 | 2020-05-18 | 225886 | 451 |
| 85 | 2020-05-19 | 226699 | 813 |
| 86 | 2020-05-20 | 227364 | 665 |
| 87 | 2020-05-21 | 228006 | 642 |
| 88 | 2020-05-22 | 228658 | 652 |
| 89 | 2020-05-23 | 229327 | 669 |
| 90 | 2020-05-24 | 229858 | 531 |
| 91 | 2020-05-25 | 230158 | 300 |
| 92 | 2020-05-26 | 230555 | 397 |
| 93 | 2020-05-27 | 231139 | 584 |
| 94 | 2020-05-28 | 231732 | 593 |
| 95 | 2020-05-29 | 232248 | 516 |
| 96 | 2020-05-30 | 232664 | 416 |
| 97 | 2020-05-31 | 233019 | 355 |
| 98 | 2020-06-01 | 233197 | 178 |
| 99 | 2020-06-02 | 233515 | 318 |
| 100 | 2020-06-03 | 233836 | 321 |
| 101 | 2020-06-04 | 234013 | 177 |
| 102 | 2020-06-05 | 234531 | 518 |
| 103 | 2020-06-06 | 234801 | 270 |
| 104 | 2020-06-07 | 234998 | 197 |
| 105 | 2020-06-08 | 235278 | 280 |
| 106 | 2020-06-09 | 235561 | 283 |
| 107 | 2020-06-10 | 235763 | 202 |

For each record in tab 1, a *k* (eq. 6) has been calculated and the corresponding value of the detected cases 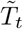 is used to synchronize the model. The model parameters are then easily found using eq. 3 with *τ* as tuning parameter. The model results and the available data are normalized to 1000 individuals.

After some trial, good *τ* values has been found within 3 and 8 days.

The next section 4 shows all results for *τ* = 7.5 *days* while in the subsection 4.1 some results for *τ* = 4 and *τ* = 8 are shown.

**Table 3:** Parameters of the model referring to Italy. Each k is calculated using the detection criteria and reported in tab. 1. The only tuning parameter is the incubation period τ. The basic reproduction number is calculated as spectral radius of the next generation matrix and in this simplified, model (see [1] and eq. 3) is equal to: 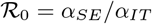

| t | Day | $\tilde{T}_t$ | $\Delta\tilde{T}_t$ | $k$ | $\tau$ | $\alpha_{EI}$ | $\alpha_{SE}$ | $\alpha_{IT}$ | $\mathcal{R}_0$ |
| --- | --- | --- | --- | --- | --- | --- | --- | --- | --- |
| 6 | 2020-03-01 | 1694 | 566 | 1.215 | 7.5 | 0.133 | 4.095 | 0.162 | 25.3 |
| 9 | 2020-03-04 | 3089 | 587 | 0.884 | 7.5 | 0.133 | 2.141 | 0.118 | 18.2 |
| 10 | 2020-03-05 | 3858 | 769 | 0.820 | 7.5 | 0.133 | 1.861 | 0.109 | 17.0 |
| 11 | 2020-03-06 | 4636 | 778 | 0.763 | 7.5 | 0.133 | 1.636 | 0.102 | 16.1 |
| 12 | 2020-03-07 | 5883 | 1247 | 0.720 | 7.5 | 0.133 | 1.479 | 0.096 | 15.4 |
| 13 | 2020-03-08 | 7375 | 1492 | 0.683 | 7.5 | 0.133 | 1.351 | 0.091 | 14.8 |
| 14 | 2020-03-09 | 9172 | 1797 | 0.650 | 7.5 | 0.133 | 1.245 | 0.087 | 14.4 |
| 16 | 2020-03-11 | 12462 | 2313 | 0.588 | 7.5 | 0.133 | 1.059 | 0.078 | 13.5 |
| 17 | 2020-03-12 | 15113 | 2651 | 0.565 | 7.5 | 0.133 | 0.995 | 0.075 | 13.2 |
| 19 | 2020-03-14 | 21157 | 3497 | 0.524 | 7.5 | 0.133 | 0.884 | 0.070 | 12.7 |
| 20 | 2020-03-15 | 24747 | 3590 | 0.505 | 7.5 | 0.133 | 0.838 | 0.067 | 12.4 |
| 23 | 2020-03-18 | 35713 | 4207 | 0.456 | 7.5 | 0.133 | 0.718 | 0.061 | 11.8 |
| 24 | 2020-03-19 | 41035 | 5322 | 0.442 | 7.5 | 0.133 | 0.688 | 0.059 | 11.7 |
| 25 | 2020-03-20 | 47021 | 5986 | 0.430 | 7.5 | 0.133 | 0.661 | 0.057 | 11.5 |
| 26 | 2020-03-21 | 53578 | 6557 | 0.419 | 7.5 | 0.133 | 0.636 | 0.056 | 11.4 |

The graphical results are shown in the following sections.

## 4 Graphic results for Italy

In all graphs the orange point represent the day where the daily differences exceeds the previous maximum daily difference. In that day an exponential parameter *κ* is calculated (see eq. 6).

The simplified SEIT model parameters have been found using the relations in (3) with the only tuning parameter *τ* corresponding to an ‘average’ incubation period. All data are normalized and refers to 1000 individuals. The detected data used to estimate the model are represented with a thick gray line ending with an orange point. The remaining data are represented with a thin grey line in order to be compared with the model forecasting (green line).

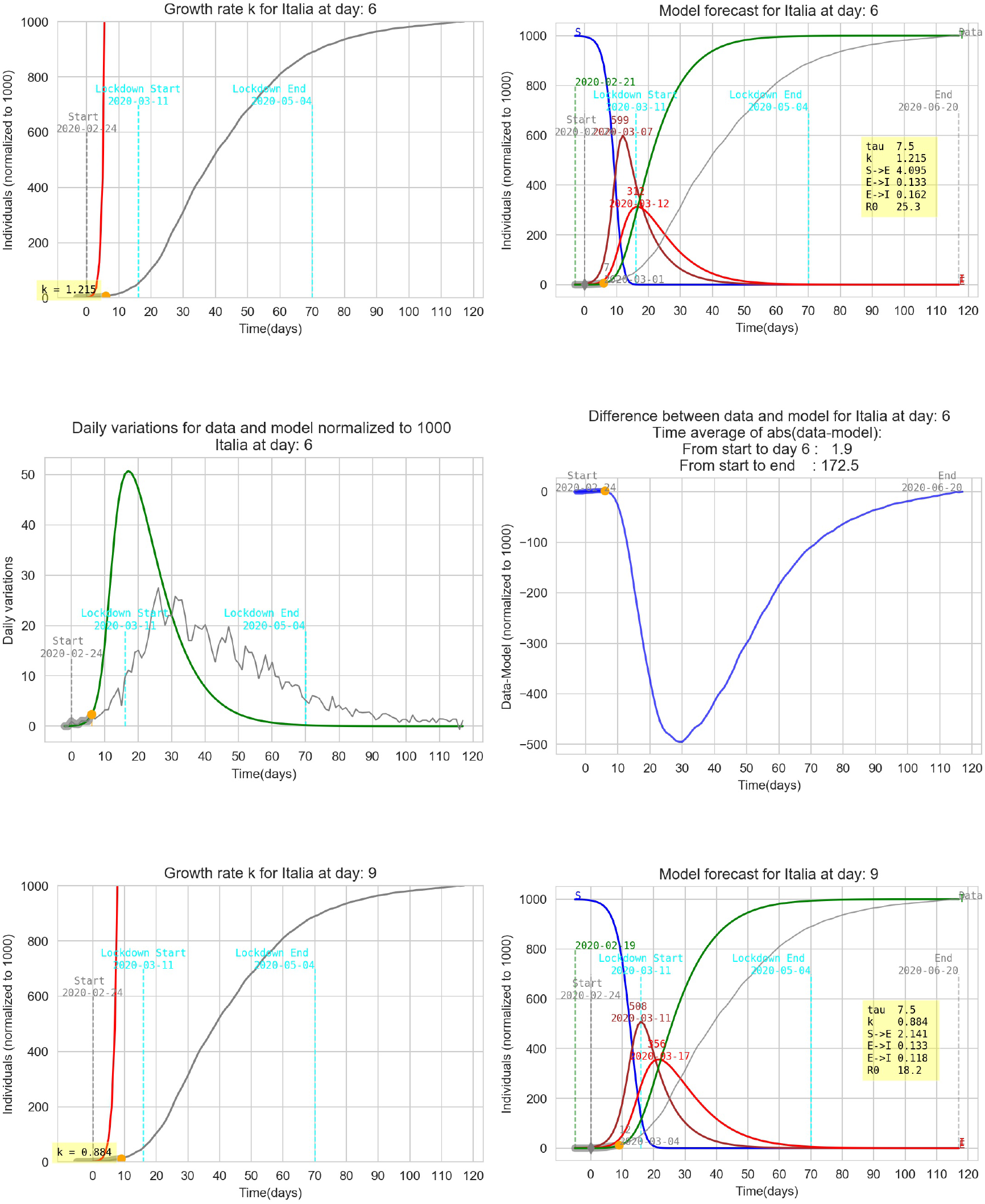

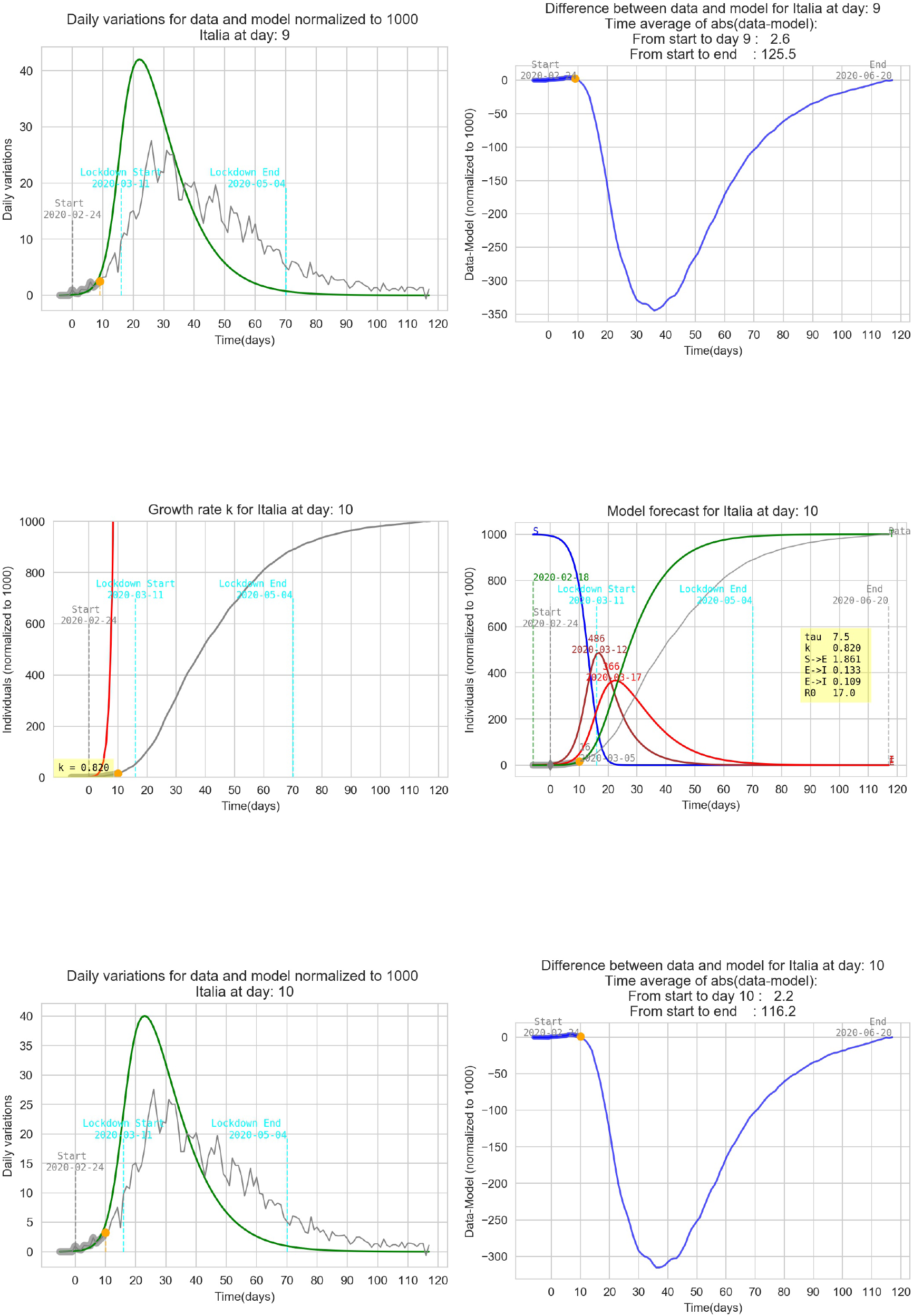

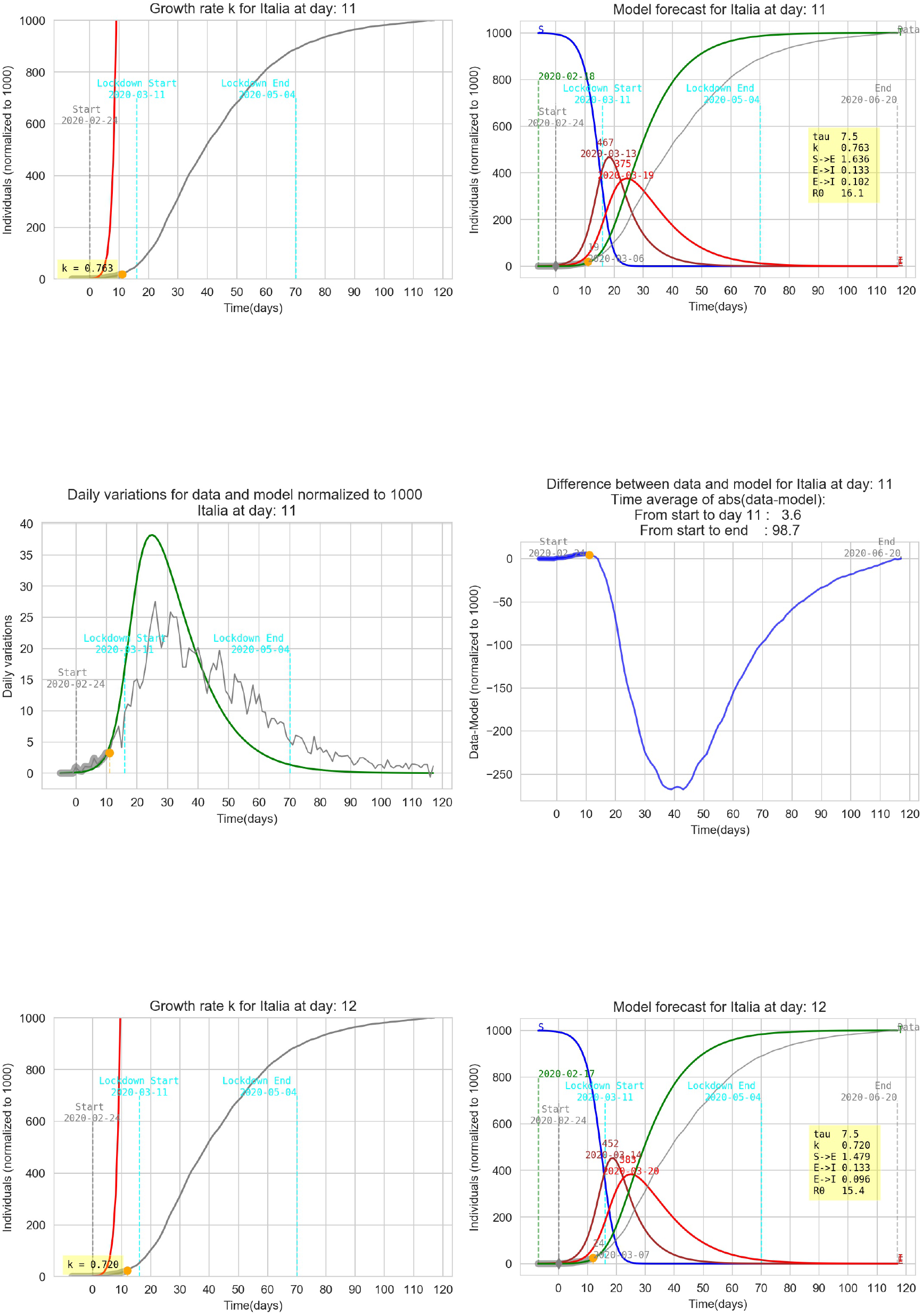

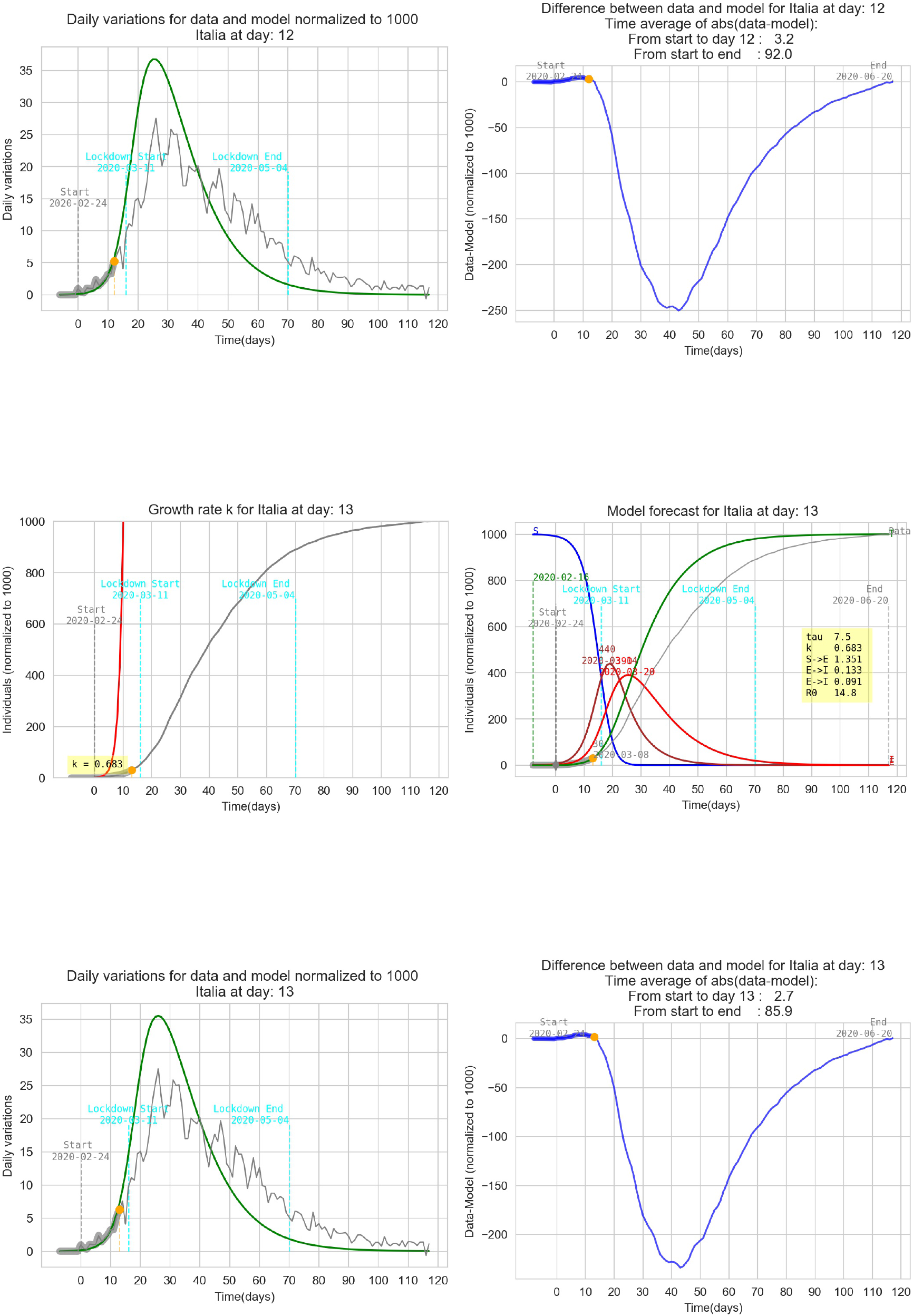

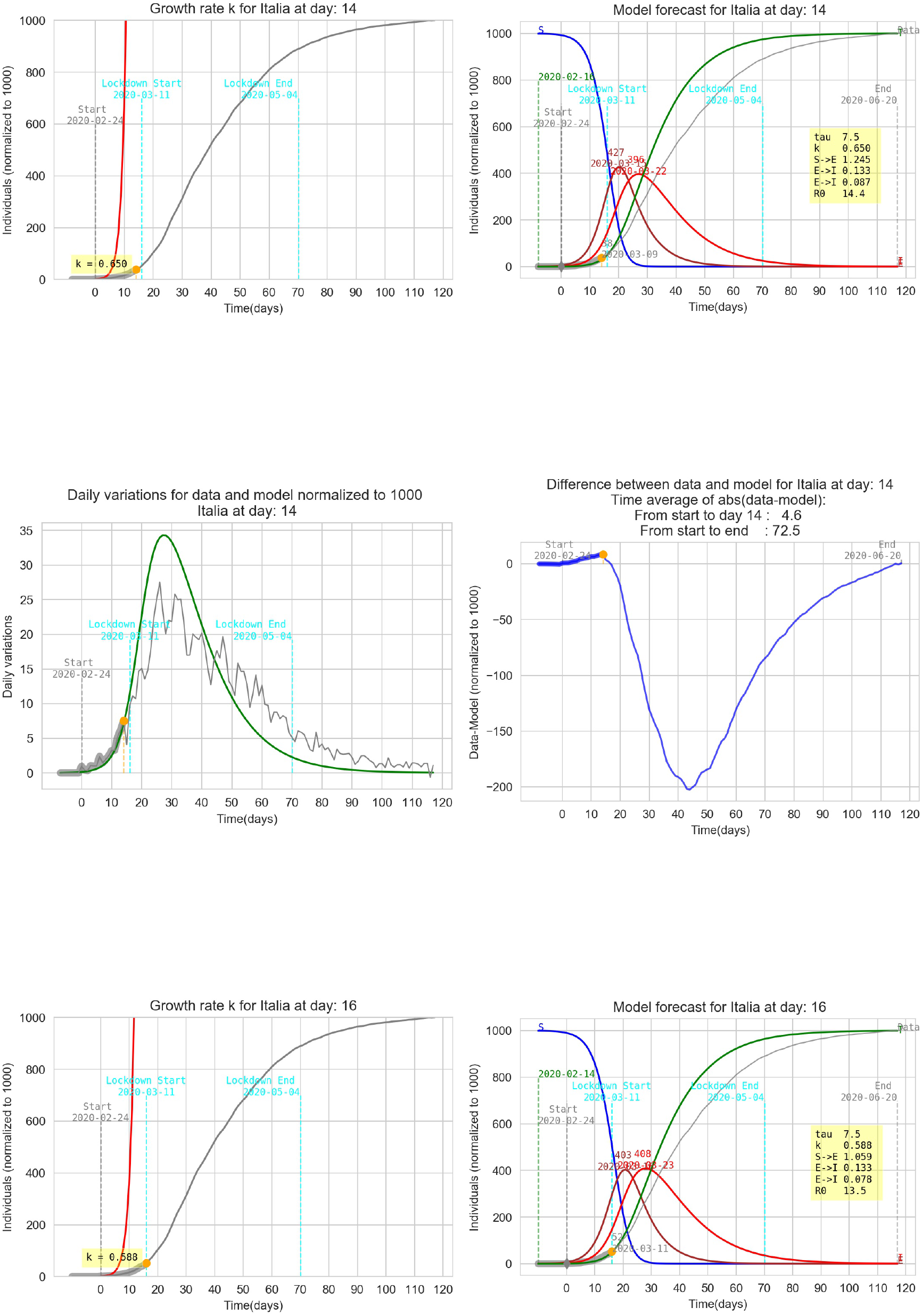

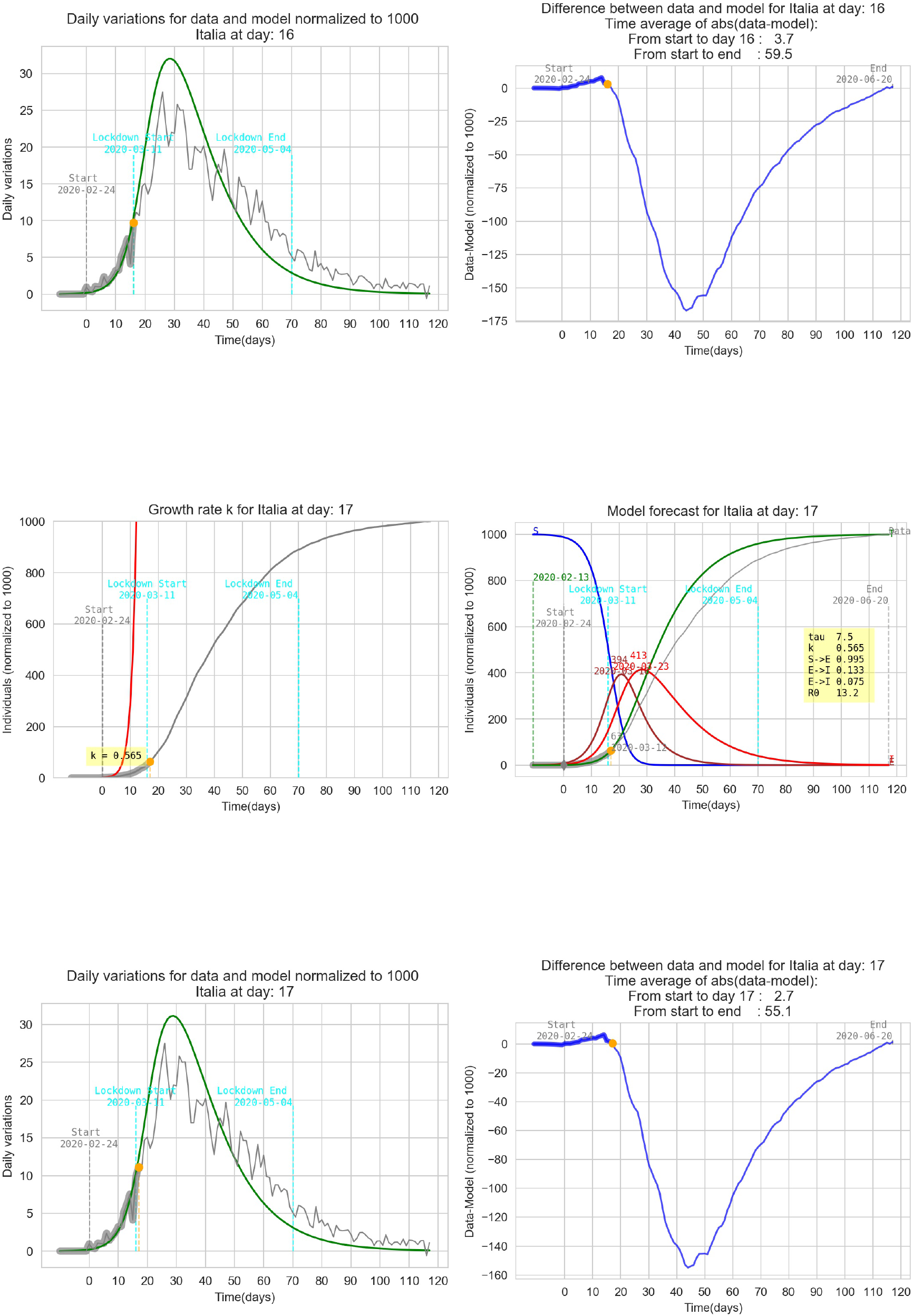

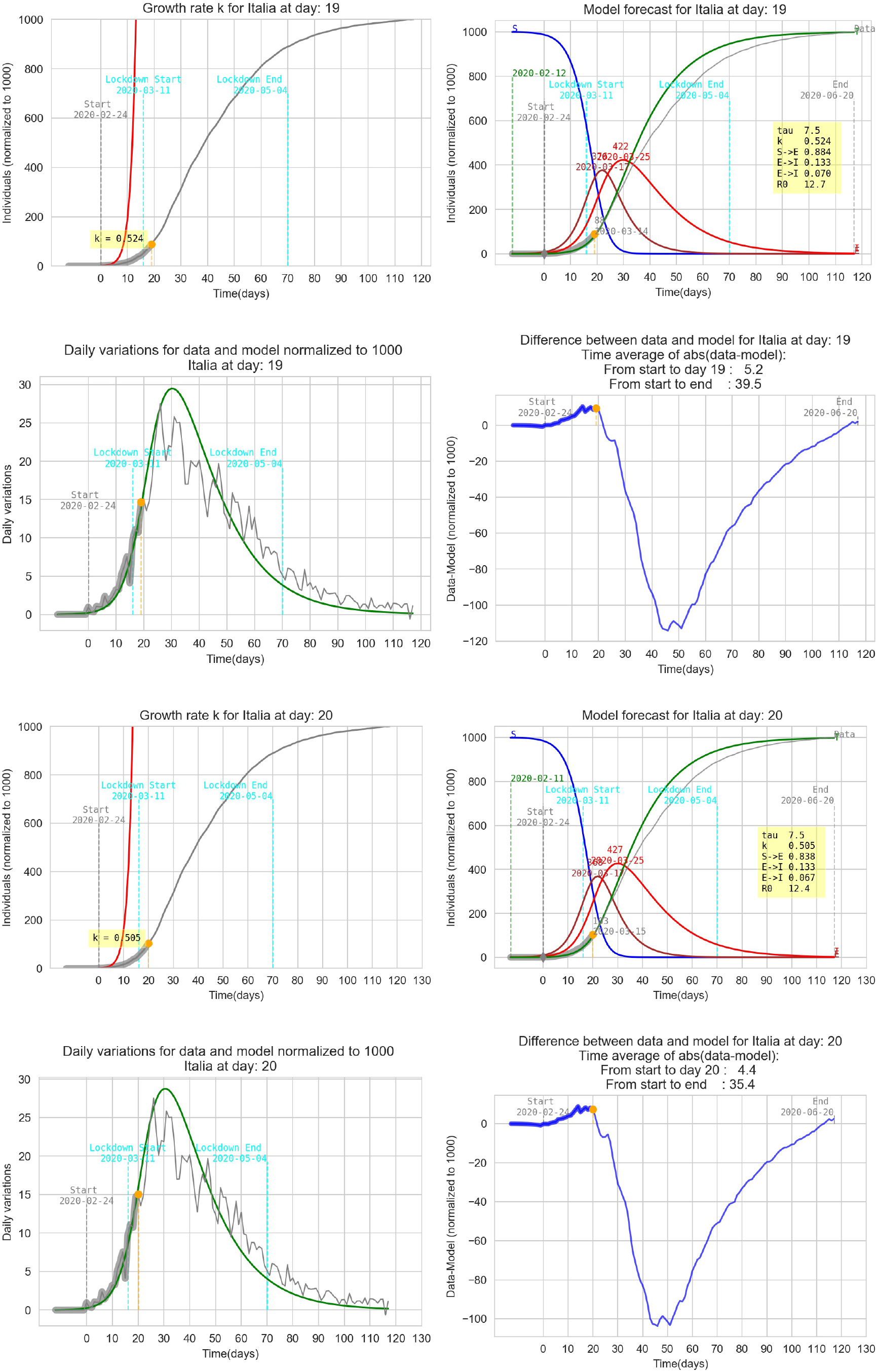

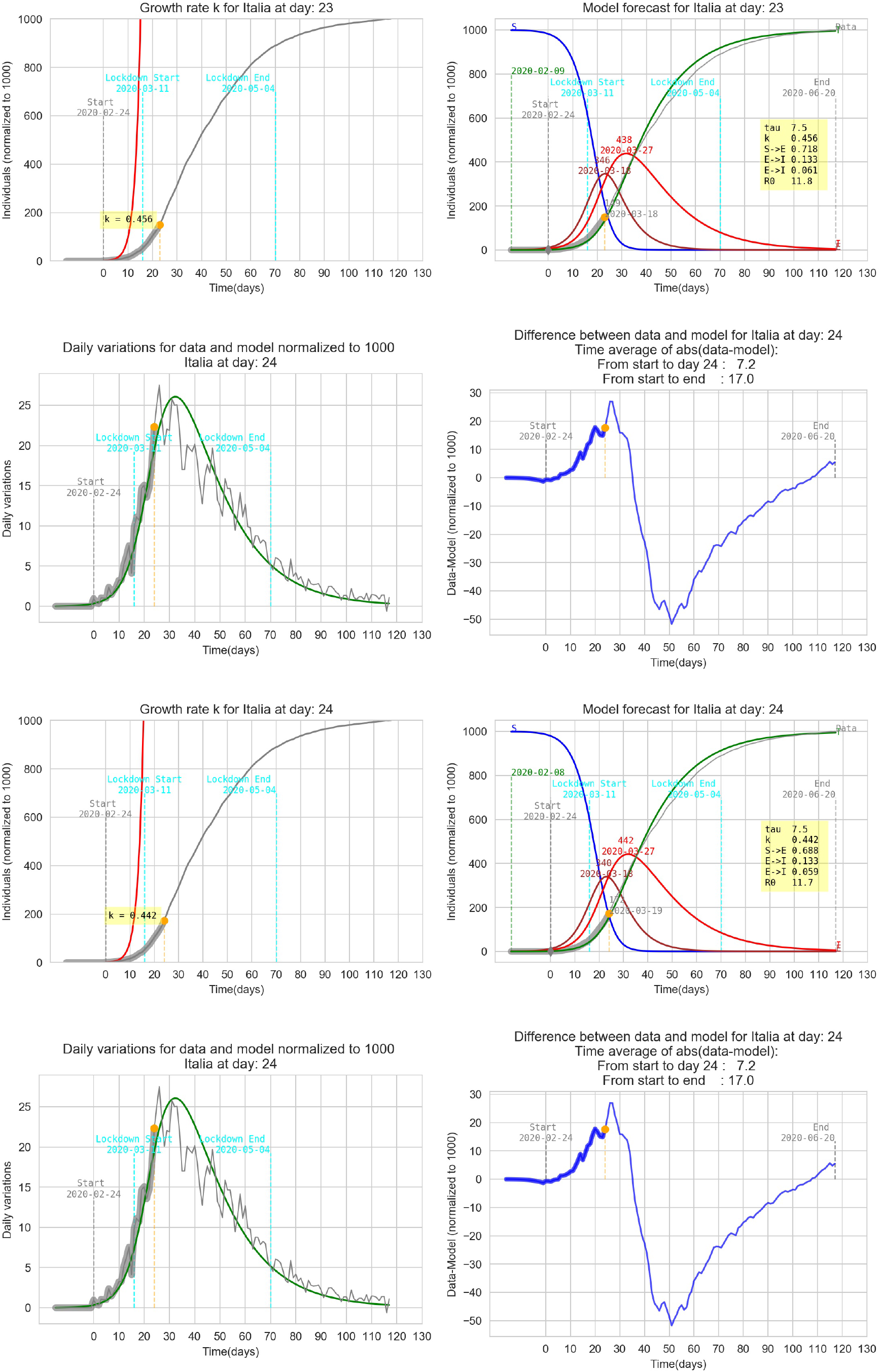

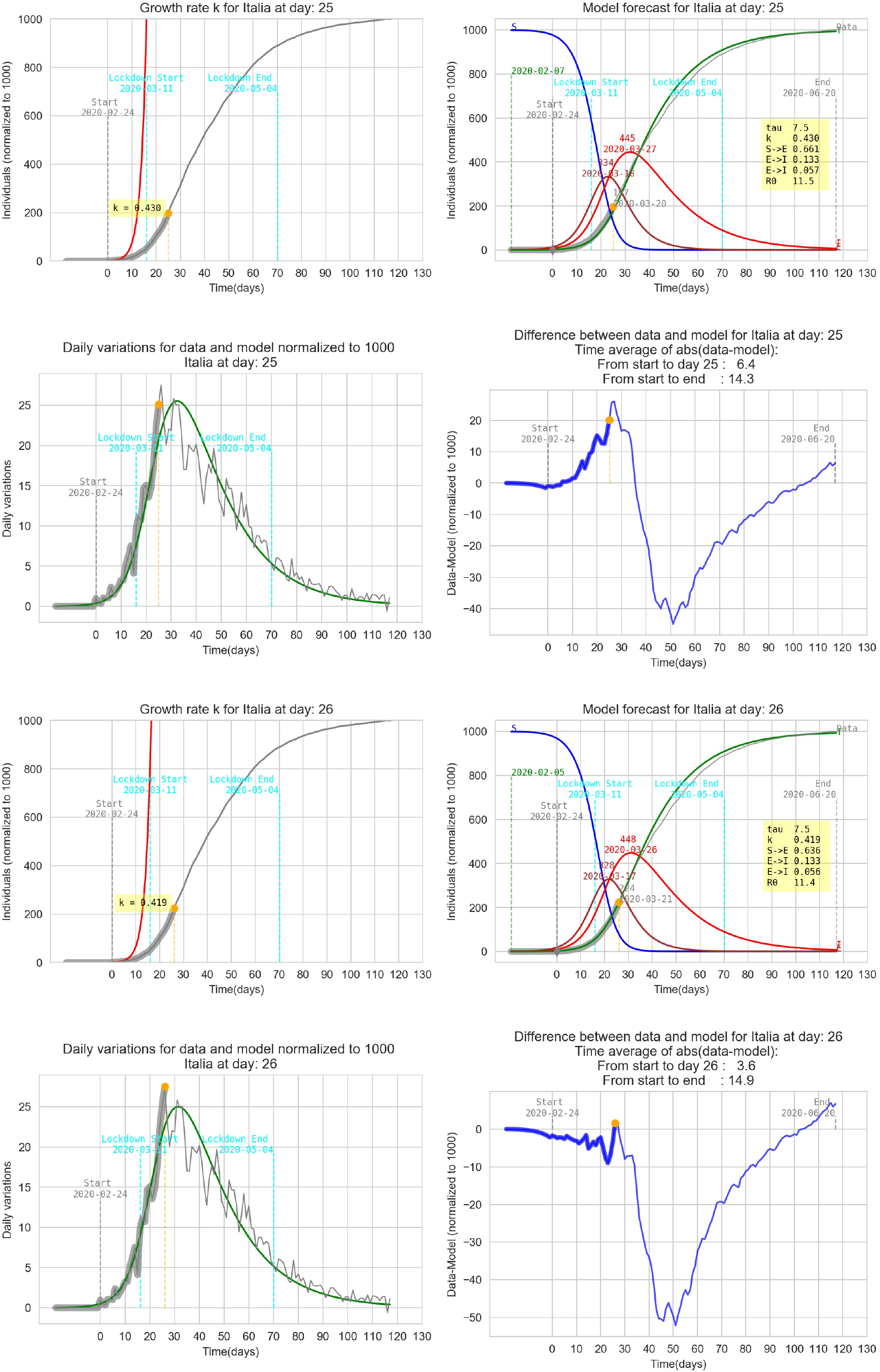

### 4.1 Sensibility to the tuning parameter

In order to have a qualitative feedback on the impact of the tuning parameter *τ* the graphs below represent the model for Italy with the detection *k* calculated at *t* = 26 and three different *τ* between 5 and 9 days. In order to have an idea of the fitting performance is calculated a time average of the absolute value of the differences between data and the model: 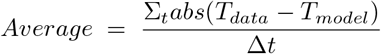.

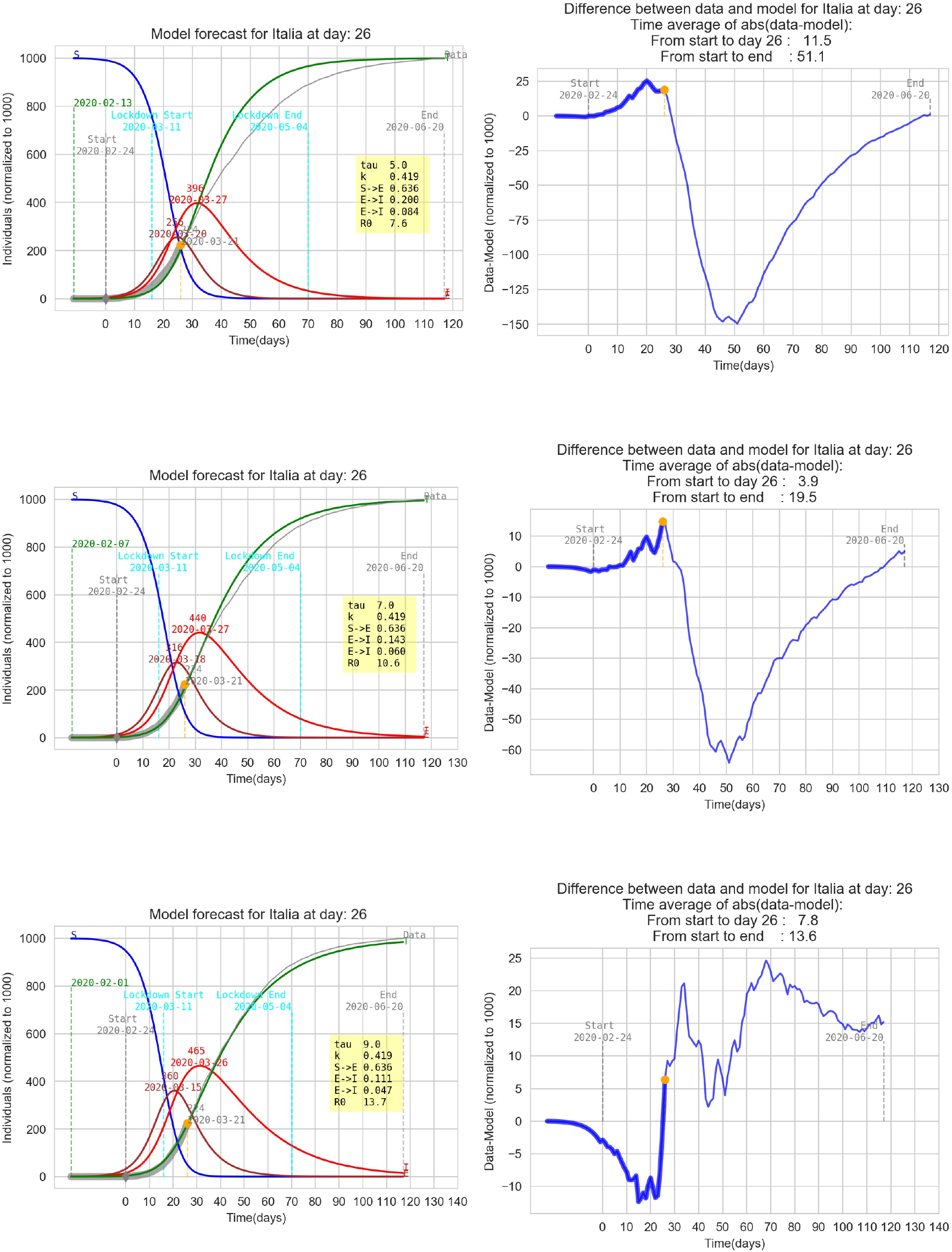

## 5 Graphic results for some italian regions

This section contains some results for several regions of Italy referred to two *k* detected in the growth phase with the same *τ*. The tuning parameter *τ* is different for each region.

### 5.1 Lombardia

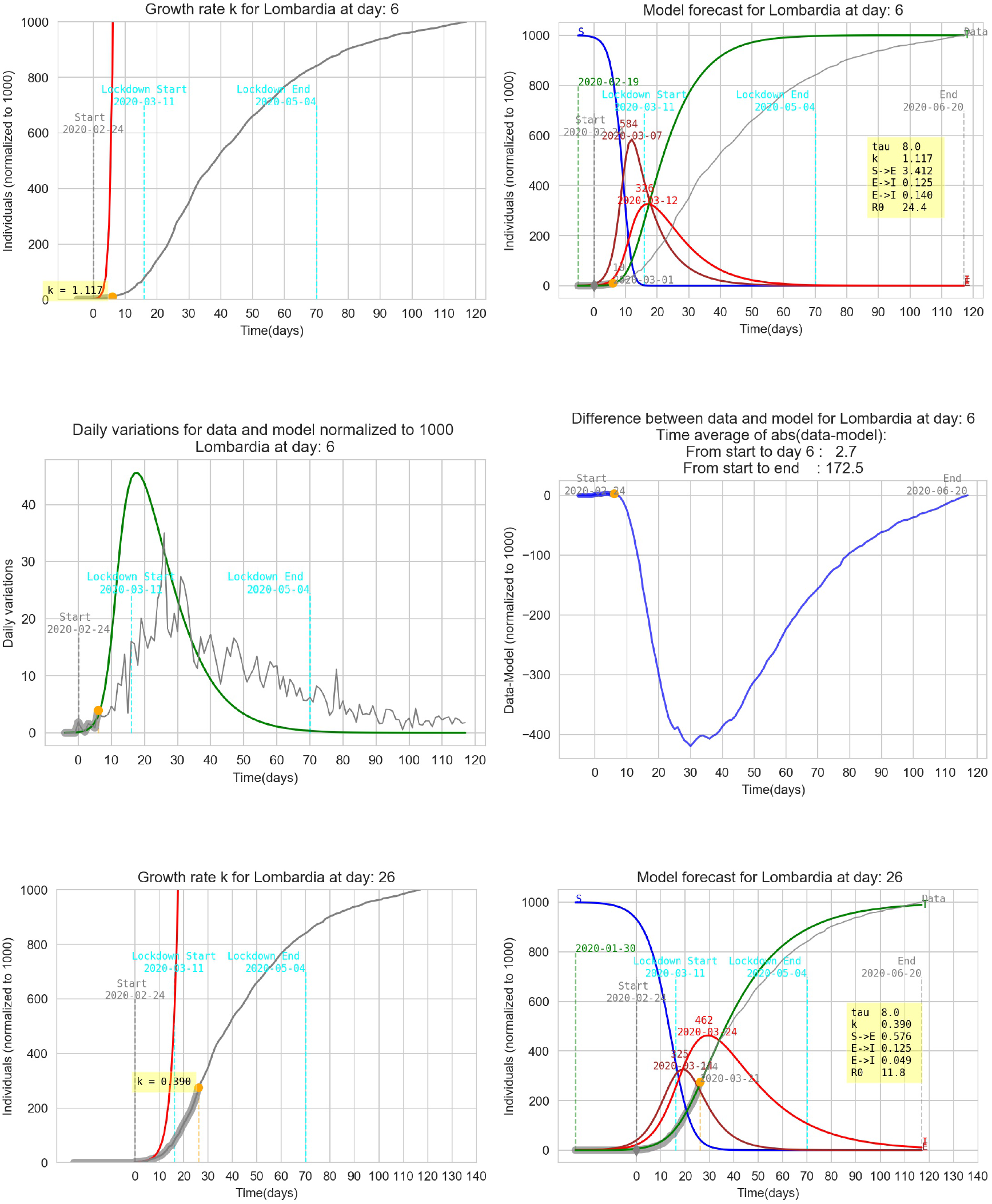

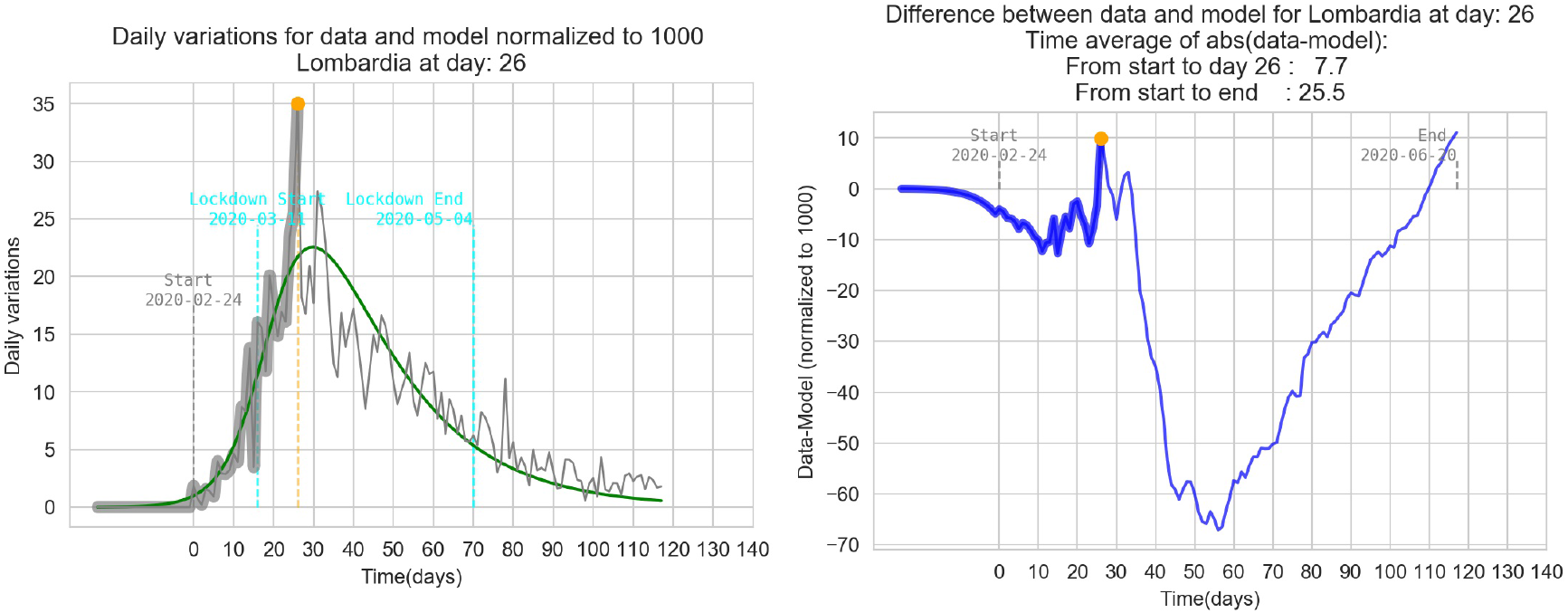

### 5.2 Piemonte

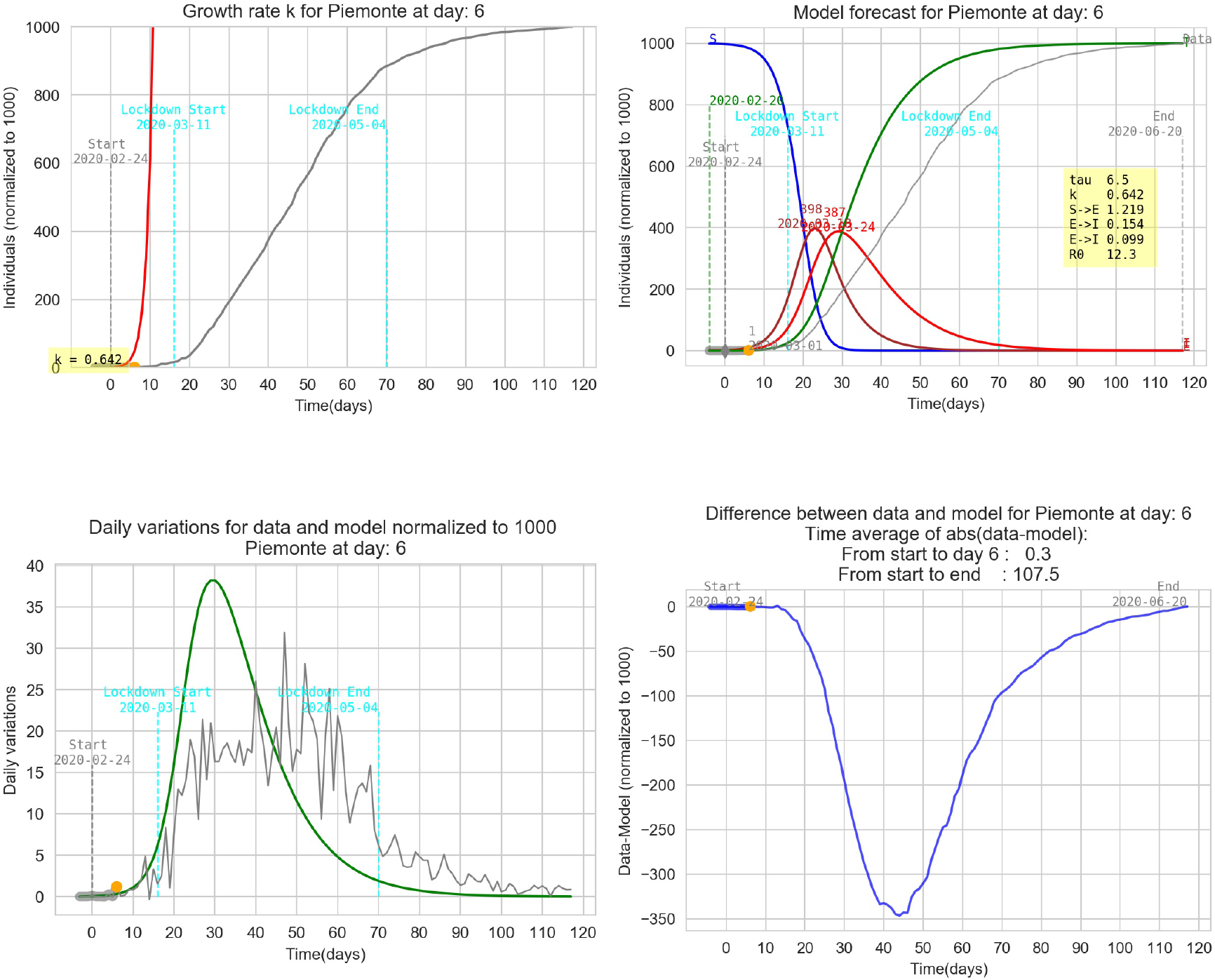

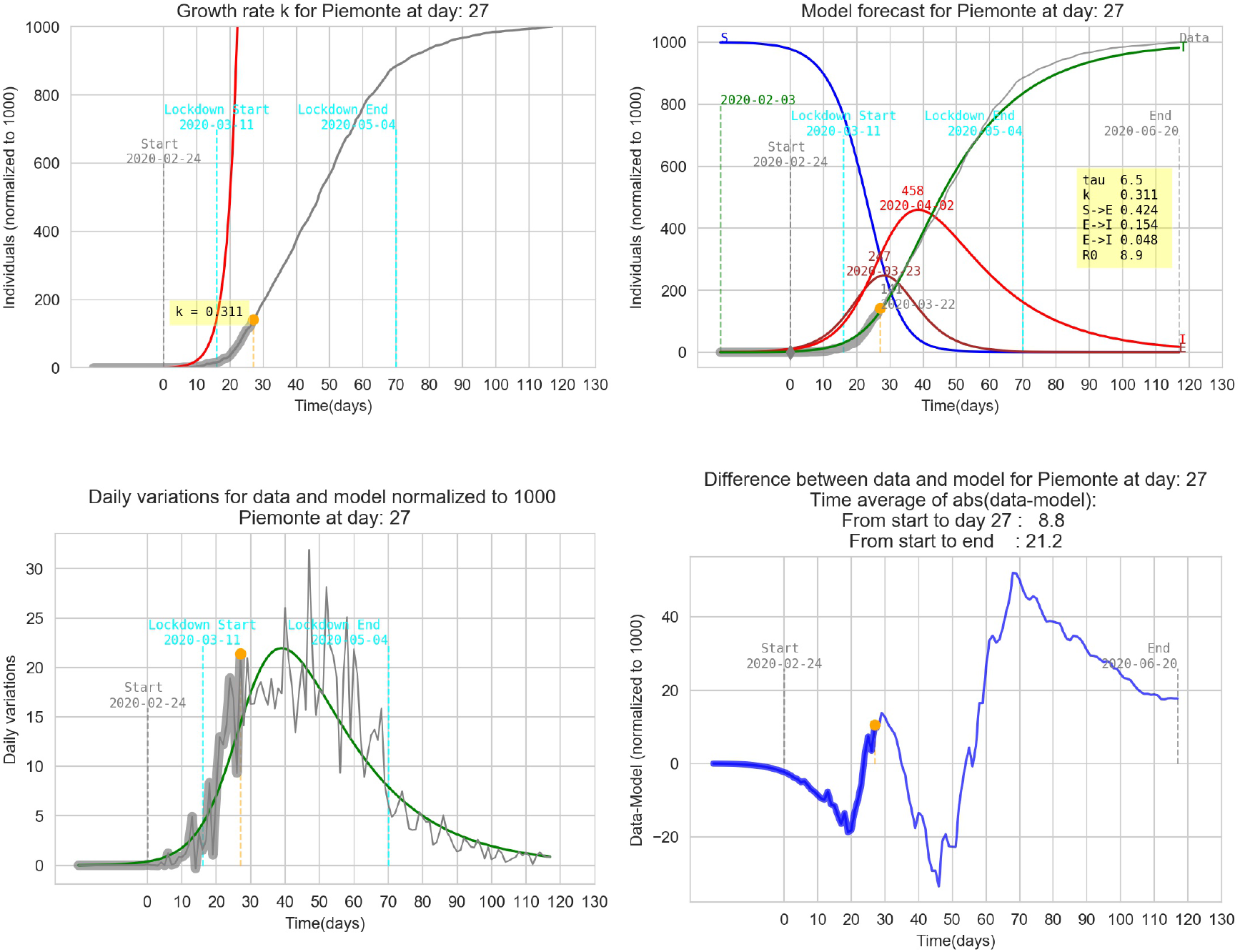

### 5.3 Toscana

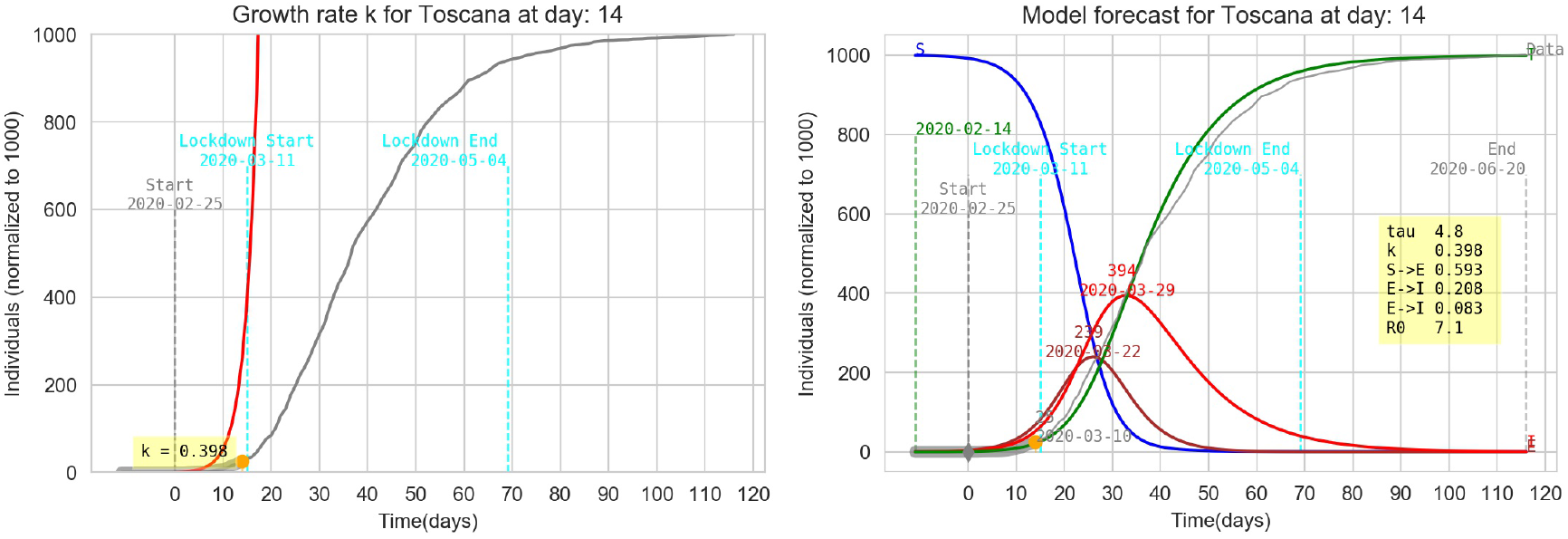

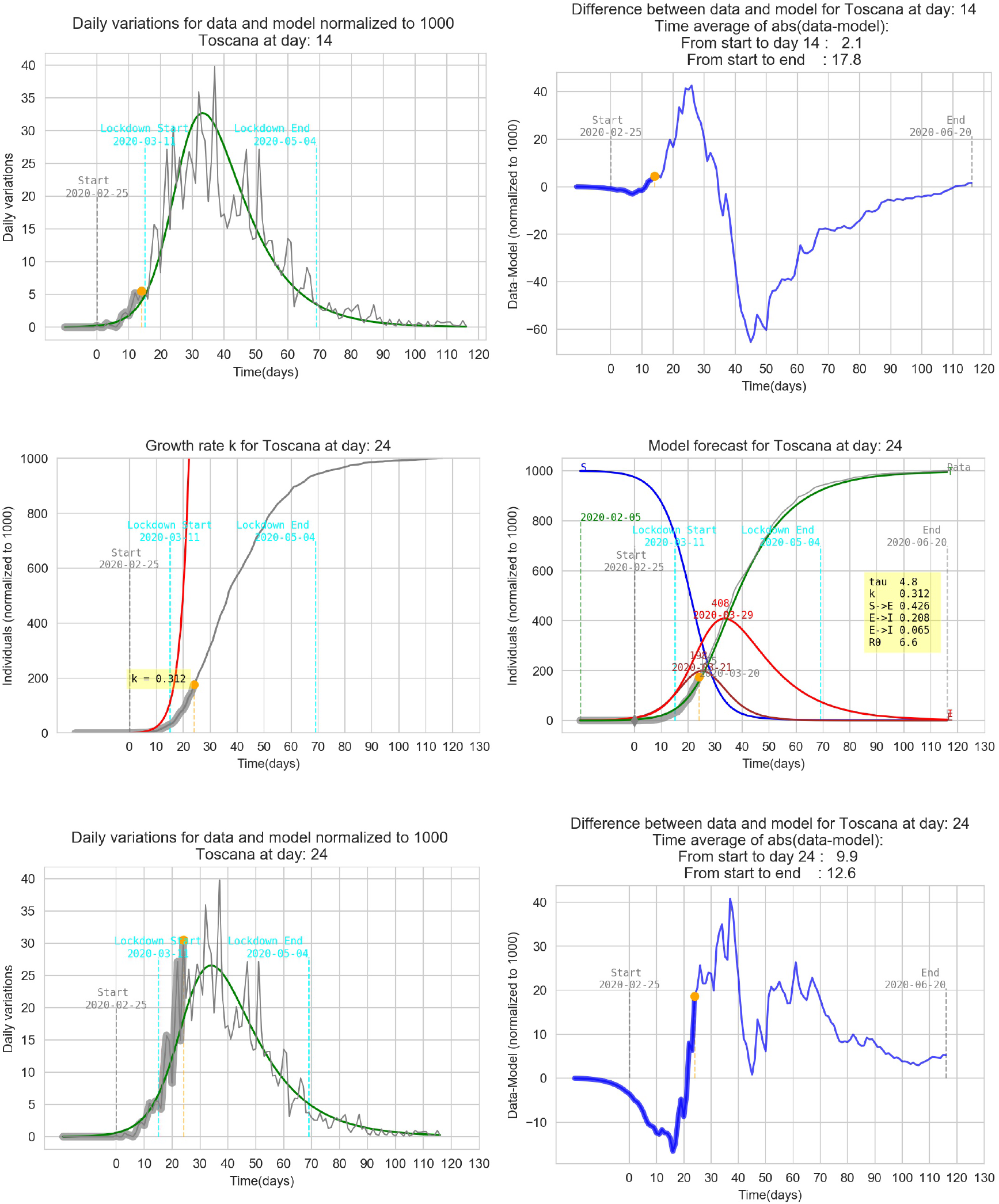

### 5.4 Lazio

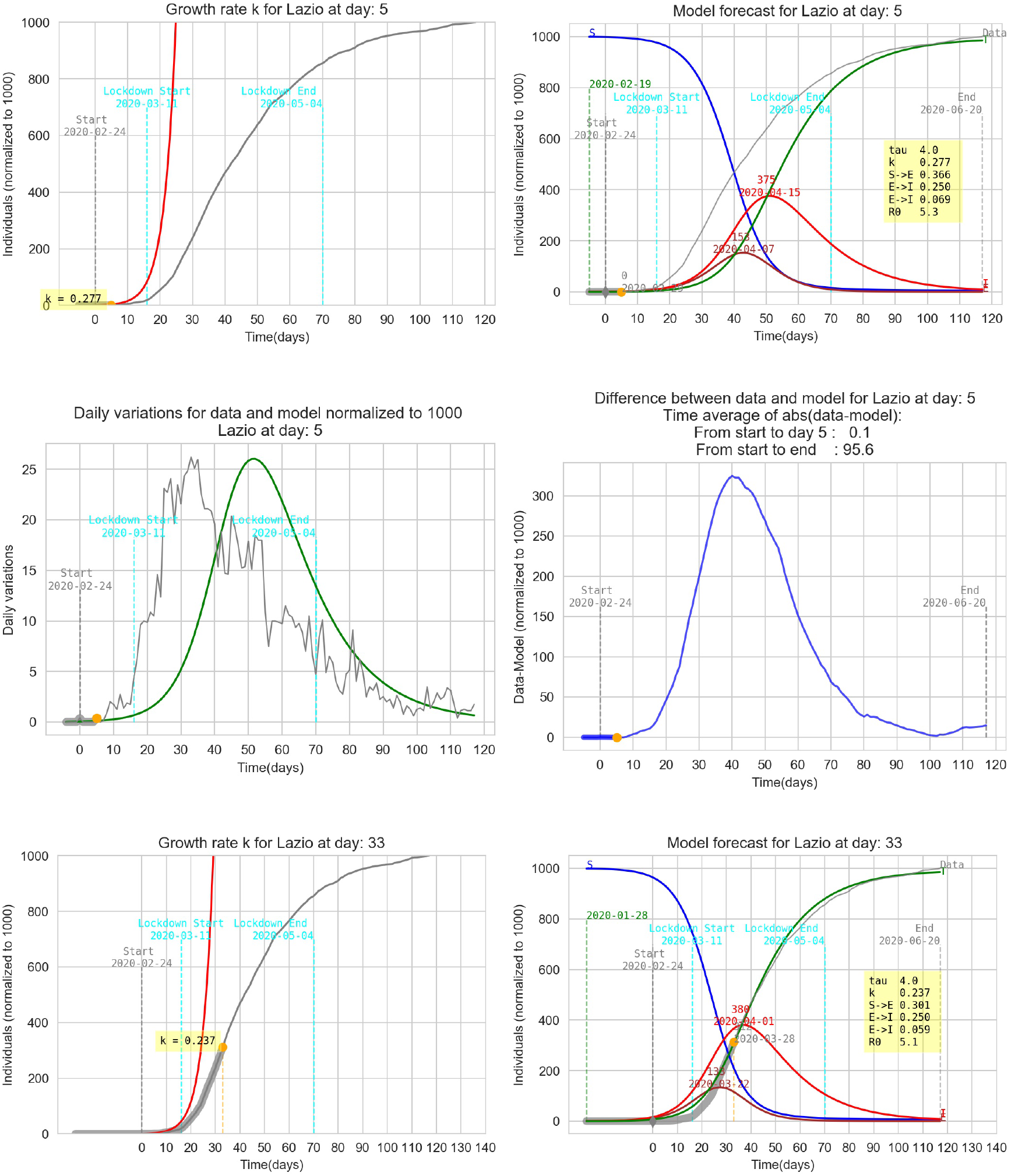

### 5.5 Pugila

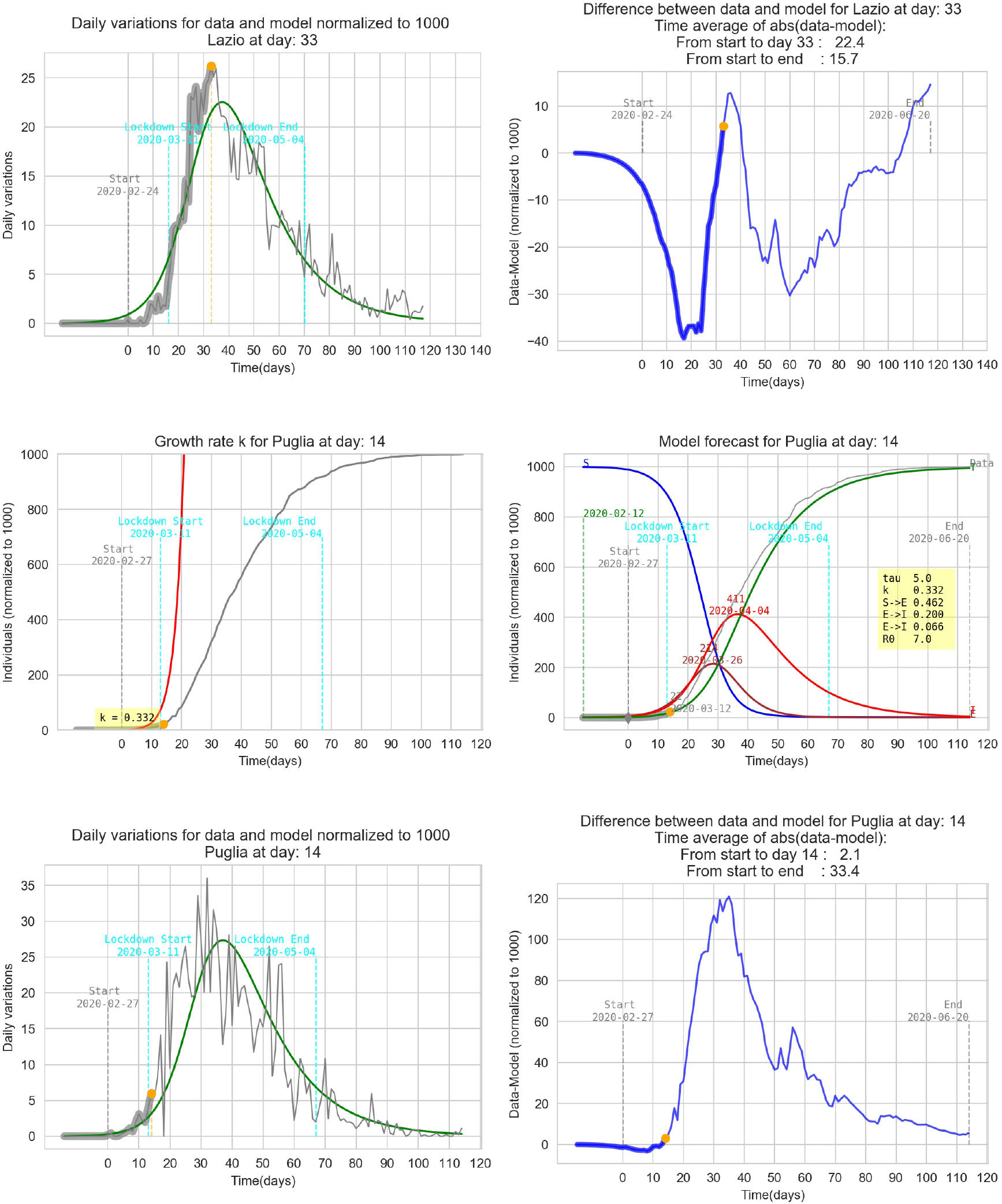

### 5.6 Calabria

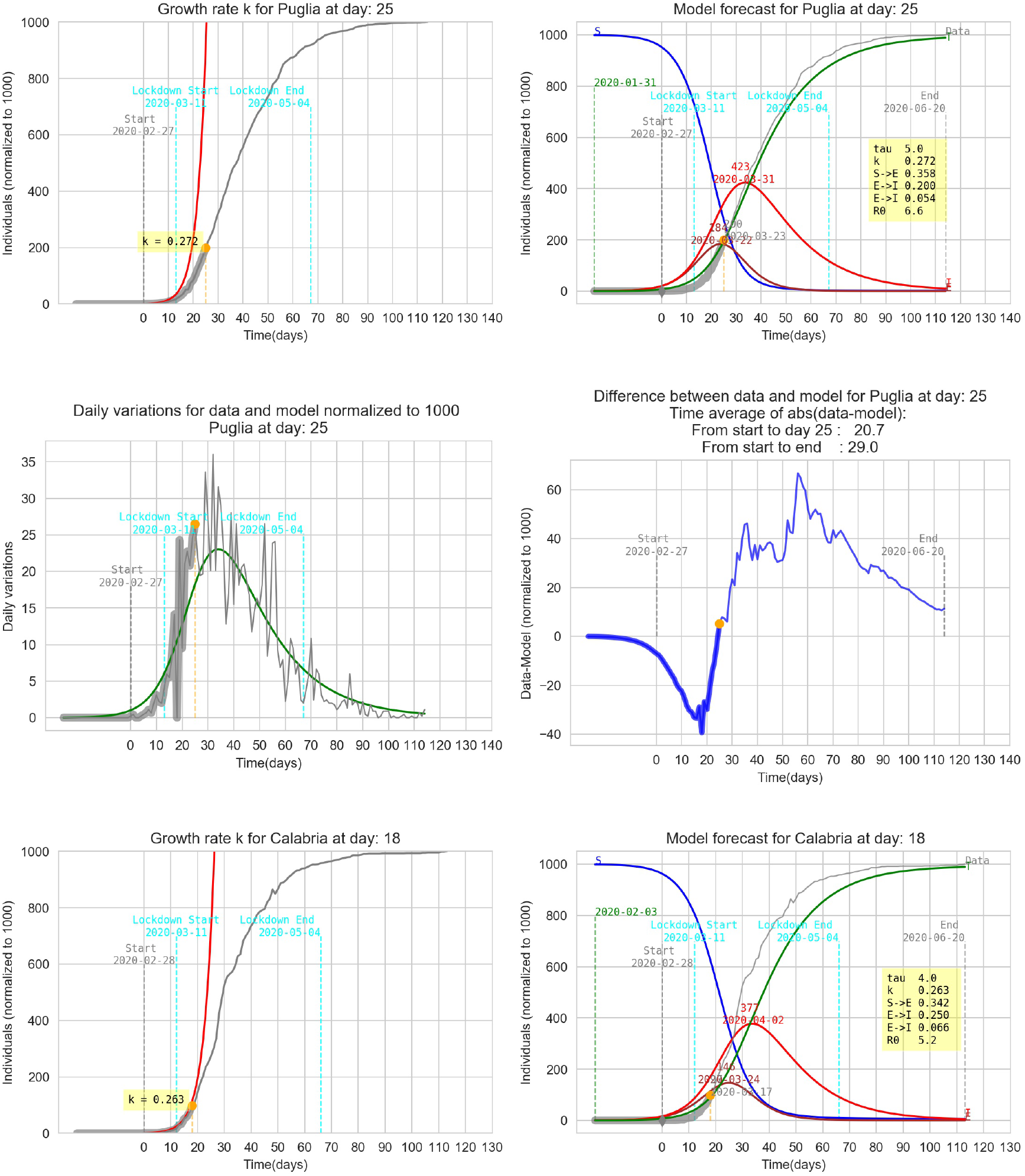

### 5.7 Sardegna

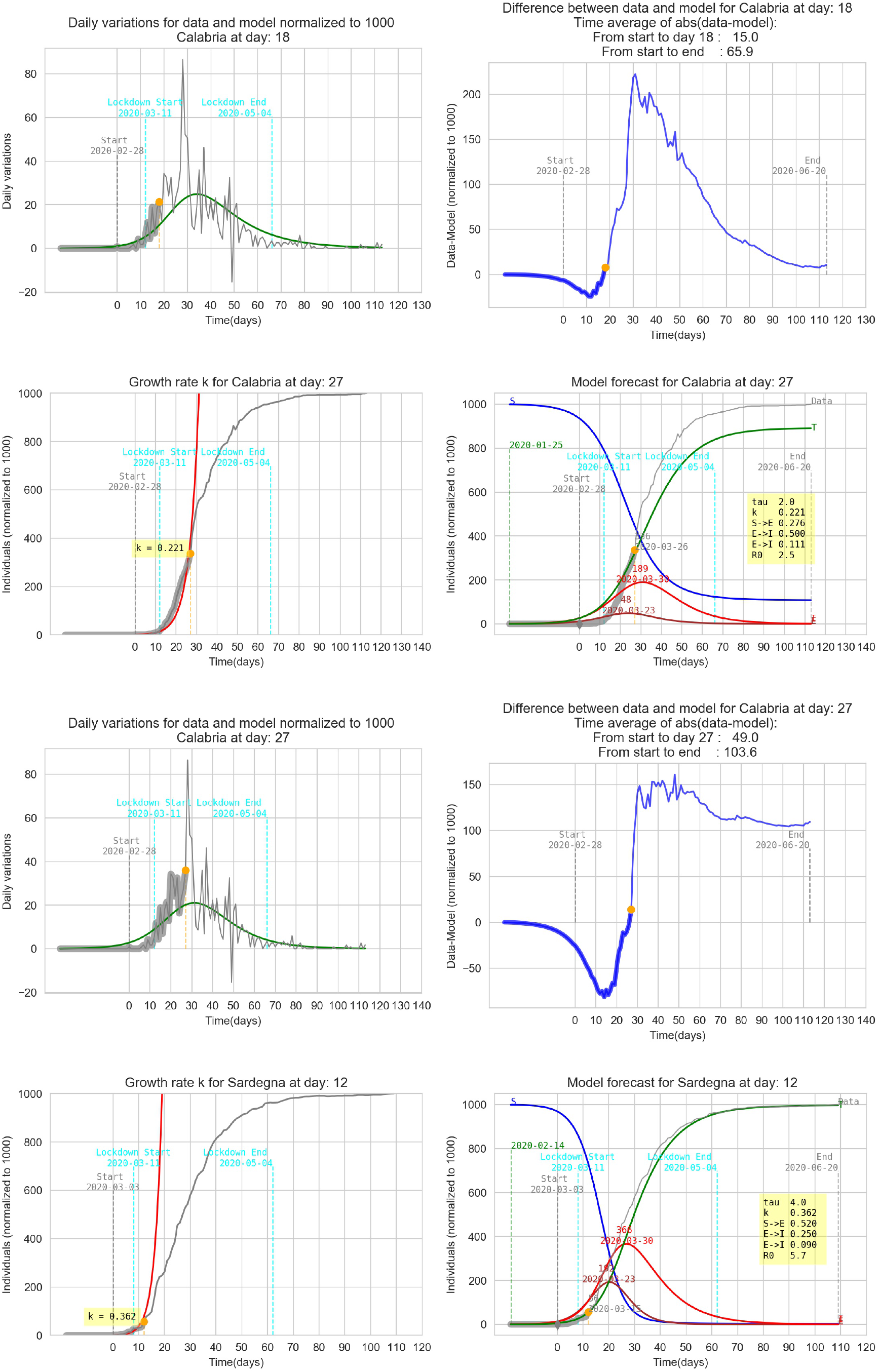

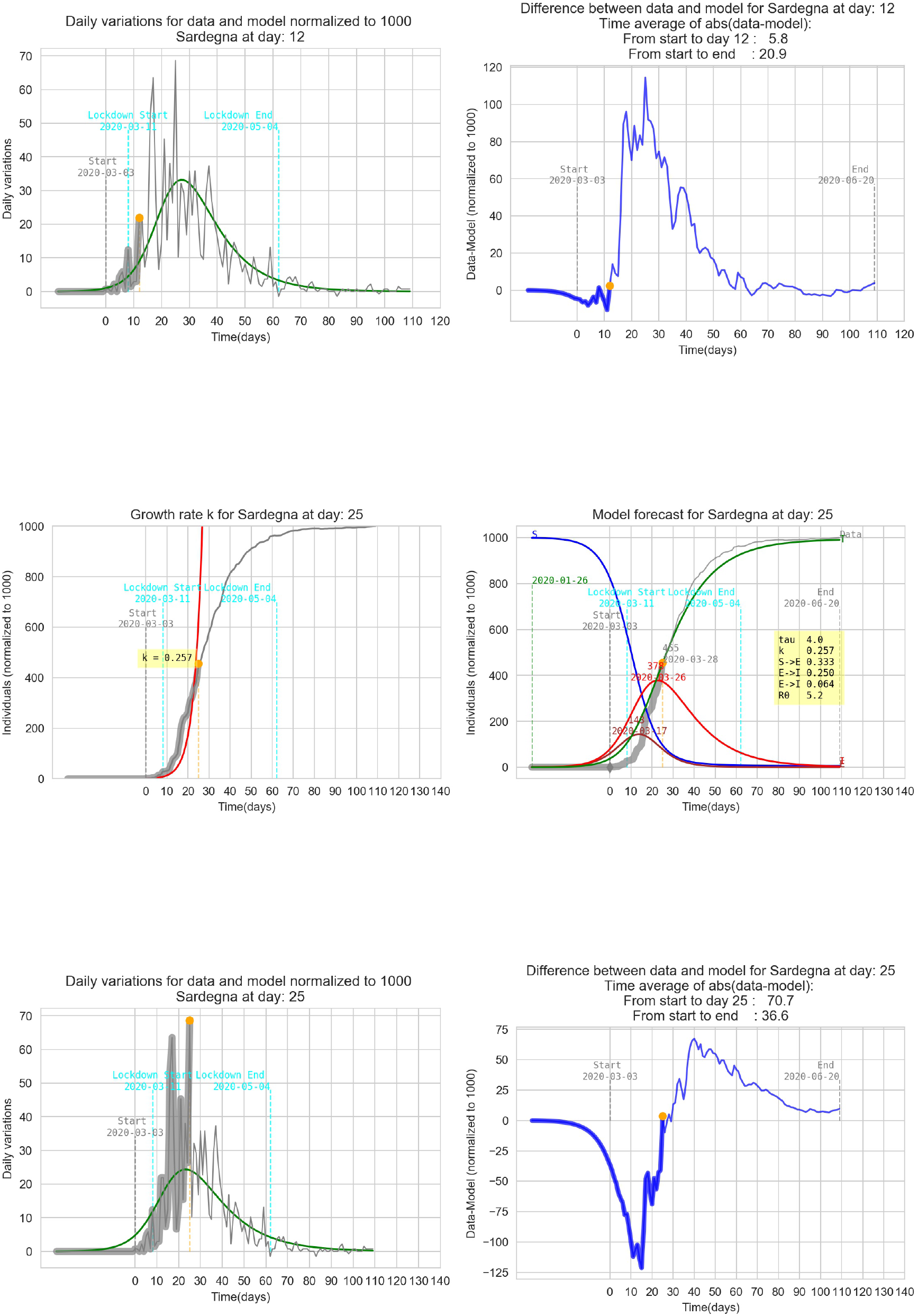

## Data Availability

the dataset used in the manuscript is publicly available on the websites managed by italian Dipartimento della Protezione Civile.

https://github.com/pcm-dpc/COVID-19/blob/master/dati-regioni/dpc-covid19-ita-regioni.csv

http://opendatadpc.maps.arcgis.com/apps/opsdashboard/index.html#/b0c68bce2cce478eaac82fe38d4138b1

